# Posttraumatic stress symptoms in chronic pain: impacts on brain morphology

**DOI:** 10.1101/2024.05.09.24307012

**Authors:** Yann Quidé, Negin Hesam-Shariati, Nell Norman-Nott, James H. McAuley, Sylvia M. Gustin

**Author notes:** **Corresponding author:** Dr Yann Quidé, NeuroRecovery Research Hub, School of Psychology, Biological Sciences (Biolink) building, Level 1, UNSW Sydney, NSW, 2052, Australia.

## Abstract

**Background:** Posttraumatic stress symptoms (PTSS) are commonly experienced by people with chronic pain. Although PTSS and chronic pain are associated with similar effects on brain morphology, the present study aimed to clarify the relationship between chronic pain and PTSS on the brain.

**Methods:** Fifty-two people with chronic pain and 38 pain-free healthy controls (HC) underwent T1-weighted magnetic resonance imaging. Severity of PTSS was measured using the civilian version of the posttraumatic stress disorder checklist (PCL-C). A series of multiple linear regressions determined the main effects of group, PTSS severity (PCL-C total score and symptom-specific scores) and their interaction on grey matter volume of selected regions-of-interest.

**Results:** The interaction term was significantly associated with variations in grey matter volume in the left and right putamen, the left middle cingulate cortex (MCC) and the right posterior insula. Results showed significantly smaller left and right putamen when reporting higher PTSS levels, and significantly larger left MCC and right posterior insula at lower PTSS levels in people with chronic pain compared to HCs. In addition, increasing PTSS severity was significantly associated with larger left and right putamen in HCs, and significantly associated with smaller left MCC and right posterior insula, in people with chronic pain.

**Conclusions:** Severity of PTSS moderated chronic pain-related grey matter alterations. More severe PTSS, especially avoidance, was associated with smaller left MCC, a core region of the “pain matrix”. The MCC is strongly linked with the somatosensory network, and critical for empathy, especially toward pain-related stimuli.

## Introduction

Chronic pain, usually defined as pain persisting for over three months (1), is a major public health concern, impacting around 28% of the global population (2). Chronic pain is a leading cause of disability and suffering (3), often associated with severe comorbid mental health problems (4), reduced quality of life (5) and increased risk for suicide (6). Exposure to traumatic events can trigger and contribute to the maintenance of pain through sensitization of the stress system, affecting stress-sensitive regions in the brain, such as the hippocampus, amygdala, the anterior cingulate cortex (ACC) or the medial prefrontal cortex (mPFC) (7). Chronic pain and trauma exposure are strongly related, with around 20% of people with chronic pain experiencing posttraumatic stress symptoms (PTSS), when posttraumatic stress disorder (PTSD) is not formally diagnosed (8), and with 20 to 80% of people with PTSD suffering from chronic pain (9). However, the relationship between chronic pain and posttraumatic stress symptoms (PTSS) is not completely understood.

The central nervous system, especially the brain, has a key role in the development and maintenance of chronic pain (10). Previous studies have highlighted chronic pain-related alterations in brain morphology, especially reduced grey matter volume in the ACC and middle cingulate cortex (MCC), the thalamus, insula, striatum, primary and secondary somatosensory cortices (11). Together, these regions have been referred to as the “pain matrix” (12, 13). A recent study reported smaller grey matter volume in the left ACC, the left anterior and posterior insula, and the left hippocampus across groups of people with migraine, craniomandibular disorder and chronic back pain, indicating these alterations were common across different chronic pain conditions and locations (14). Importantly, alterations in these key brain regions, crucial for emotion regulation and social cognition (15, 16), have been associated with comorbid mental health conditions, including depression, anxiety and PTSD (17–20). However, despite the high comorbidity between PTSD and chronic pain, previous studies have focused on identifying the functional substrates of pain in PTSD (e.g., (21, 22)), and the effects of PTSS on brain morphology in people with chronic pain conditions remains largely unknown.

Perhaps unsurprisingly, grey matter alterations in similar regions have been reported in independent studies of PTSD and chronic pain (7). The most consistent results from studies investigating PTSD, demonstrate smaller volumes of the hippocampus, ACC, insula, striatum, amygdala, middle frontal gyrus (MFG, part of the dorsolateral prefrontal cortex), and orbitofrontal/mPFC (23–27). In addition, alterations in specific brain regions have been associated with specific PTSD symptoms, with insular function being associated with hyper-arousal and re-experiencing symptoms (28, 29), while decreased ACC and increased insular function were associated with avoidance (30, 31). Despite clear morphological and functional differences (32), only a few studies have separately investigated the anterior and posterior insulae in the context of trauma or PTSD research (33). The posterior insula is highly connected to the limbic system, including the ACC, and receives interoceptive (e.g., visceral, homeostatic) and exteroceptive (e.g., sensory) information which is integrated by the anterior insula (34, 35). However, the effects of trauma on the morphological integrity of these regions in people with chronic pain, is to our knowledge largely unknown.

The present study aims to clarify the relationship between PTSS severity, chronic pain and brain morphology in key regions commonly reported in separate studies of PTSD and chronic pain, including the hippocampus, amygdala, striatum, thalamus, mPFC, MFG, ACC, MCC, and the anterior and posterior insulae. We hypothesized that grey matter volume reduction observed in chronic pain, relative to healthy controls would be dependent of the severity of PTSS reported. More pronounced grey matter volume reduction in these regions would be evident for those reporting more severe PTSS. In addition, we hypothesized that people with chronic pain would exhibit greater reductions in grey matter volume in these key brain regions in association with PTSS severity, compared to pain-free healthy controls.

## Methods and Materials

### Participants

Participants were 52 people reporting chronic pain conditions lasting for more than three months (together referred to as the *chronic pain* group), including temporomandibular disorder (n=15), trigeminal neuropathic pain (n=13), burning mouth (n=1), trigeminal neuralgia (n=6), temporomandibular disorder + trigeminal neuropathic pain (n=1), and spinal cord injury neuropathic pain (n=16; complete paraplegia with continuous burning and/or shooting pain in areas of sensorimotor loss), as well as 38 pain-free healthy controls (HC). Inclusion criteria for all participants were age over 18 years old with no known diagnosis of psychiatric disorder. Exclusion criteria included having a heart pacemaker, metal implants, intrauterine contraceptive device, insulin pump, infusion devices, hearing-disease, claustrophobia, pregnancy, a history of stroke, multiple sclerosis, or Parkinson’s disease. All participants were volunteers who provided informed consent according to procedures approved by the Human Research Ethics committees of the University of New South Wales (HC15206), the University of Sydney (HREC06287) and Northern Sydney Local Health District (1102-066M).

### Assessments

The civilian version of the PTSD Checklist (PCL-C) (36) is a standardized self-report 17-item questionnaire used to measure the severity and burden of PTSS. Participants indicate how much they have been bothered by a symptom over the past month using a 5-point scale (1 = not at all, 5 = extremely). In this study, no provision of a formal PTSD diagnosis was intended. The overall burden of PTSS (PCL-C total score, ranging from 17 to 85) was first established in focal analyses, followed by scoring for specific PTSS including re-experiencing (cluster B; questions 1-5), avoidance (cluster C; questions 6-12) and hyperarousal symptoms (cluster D; questions 13-17). Severity of depressive symptoms was measured using the sum of all 21 items from the Beck Depression Inventory (BDI-I total scores ranging from 0 to 63) (37), and the severity of state anxiety was assessed using the 20-item of the State subscale (scores ranging from 20 to 80) from the State-Trait Anxiety Inventory (STAI) (38).

A visual analogue scale (VAS) was used to evaluate participant’s pain intensity in two ways. First, participants reported their experienced levels of pain on a 10-cm horizontal ruler (‘no pain’ = 0 cm mark; ‘worst pain imaginable’ = 10 cm mark) three times a day (morning, noon, and evening) during the week (7 days) preceding their visit to the scanner (the ‘pain diary’). Second, pain during the scanning session was rated as soon as the participant left the scanner (the ‘scan pain’) (Table 1).

**Table 1.**
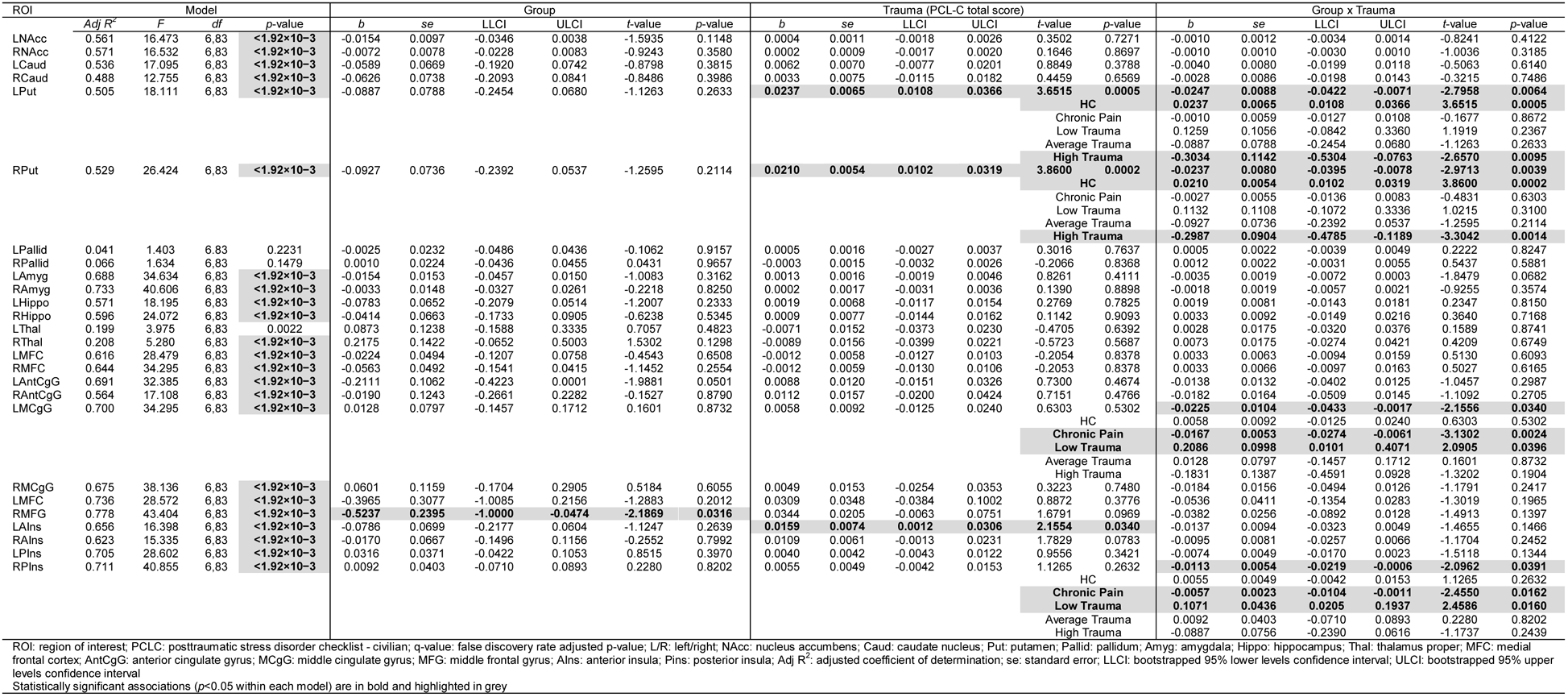
Sociodemographic and clinical characteristics of the studied cohort.

### Magnetic resonance imaging

Imaging data were acquired for each participant on Philips 3T Achieva TX scanners (Philips Healthcare, Best, The Netherlands) housed at Neuroscience Research Australia (Randwick, New South Wales, Australia; HC, n = 24, Chronic Pain, n = 14) or at St Vincent’s Hospital (Darlinghurst, New South Wales, Australia; HC, n=39, Chronic Pain, n=13). Both scanners were equipped with eight-channel head-coils and used the same acquisition parameters to collect 3D T1-weighted MPRAGE structural scans (repetition time = 5.6 ms, echo time = 2.5 ms, field of view = 250 × 250 × 174 mm, matrix 288 × 288, 200 sagittal slices, flip angle = 8°, voxel size 0.9 × 0.9 × 0.9 mm).

A radiologist reviewed all scans before releasing them to the study investigators. An additional visual inspection for gross artefacts and movements (presence of excessive ringing that would not allow identification of two adjacent brain regions) was followed by an automated quality control using the Computational Anatomy Toolbox (CAT12.8.1_2043; http://dbm.neuro.uni-jena.de/cat/index.html) for SPM12 (v7771; Wellcome Trust Centre for Neuroimaging, London, UK; http://www.fil.ion.ucl.ac.uk/spm) in MATLAB r2022a (Mathworks Inc., Sherborn, MA, USA). Structural scans were pre-processed using CAT12 default routine for voxel-based morphometry (VBM; see https://neuro-jena.github.io/cat12-help/#process_details). Following these steps, an additional quality control on sample homogeneity was performed to ensure there were no outlier scans (Mahalanobis distance between mean correlations and weighted overall image quality significantly higher than the other scans). Grey matter (GM) images were smoothed with an 8 mm full width at half maximum Gaussian kernel. Finally, total intracranial (TIV), total GM, total white matter (WM) and total cerebrospinal fluid (CSF) volumes were extracted for each participant. Average grey matter volumes for the selected 26 regions of interest (ROIs; left and right hippocampus, amygdala, caudate, putamen, pallidum, nucleus accumbens, thalamus, mPFC, MFG, ACC, MCC, anterior and posterior insula) were extracted from the Neuromorphometrics atlas in CAT12 for focal analyses.

### Harmonization

Before conducting statistical analyses, individual pre-processed images and extracted ROI values were harmonized using the python-based neuroHarmonize tools (https://github.com/rpomponio/neuroHarmonize) (39). This approach uses empirical Bayes methods derived from the ‘ComBat’ R package (40) to adjust whole-brain statistical maps and MRI-derived indices of brain morphometry (GMV, WMV, CSF, TIV and ROIs) for variations associated with scanning location in multi-site MRI studies. Age, sex, group, and PCL-C total scores were modelled as covariates during harmonization to ensure neuroHarmonize does not remove the variance associated with these variables.

### Statistical analyses

A series of multiple linear regressions were performed to determine the main effects of group (HC versus chronic pain), severity of PTSS (PCL-C total score) and their interaction (the product of group × mean-centered PCL-C total score), first on grey matter volume of each a-priori ROI (one model for each ROI), and second on whole-brain VBM maps. Age, sex and (harmonized) TIV were added as covariates in all neuroimaging analyses.

For ROI analyses, only models surviving Bonferroni correction to account for the number of ROIs tested were considered (*p*=0.05/26=1.92×10^−3^). For the whole-brain analysis in SPM12/CAT12, statistical significance was set at an initial uncorrected voxel-wise threshold of *p*<0.001, to which family-wise error correction was applied to the cluster statistics (family-wise error-corrected *p*-threshold (*pFWEc*)<0.05). When significant effects were detected, raw signal at the cluster peak was extracted for further analyses in R (v4.3.1) (41) and RStudio (2023.6.2.561) (42).

In case of significant interactions, moderation analyses were performed using the ‘interactions’ R package (v1.1.5) (43). Two sets of moderation analyses were performed separately on each ROI or on the extracted raw signal at the cluster peak as the dependent variable. In the first moderation analysis, the effects of group (independent variable) were tested at three levels of PTSS severity (PCL-C total score; moderator): at 1 standard deviation (SD) below the average PCL-C total score (low PCL-C total score), at average PCL-C total score, and at 1 SD above the average PCL-C total score (high PCL-C total score) (44). In the second moderation analysis, the effects of PCL-C total score (independent variable) on indices of grey matter volume were tested for each group (moderator). The Davidson–MacKinnon correction (HC3) was used to account for potential issues related to heteroskedasticity (45) using the R package ‘sandwich’ (v3.2.2) (46, 47). Within each significant model, statistical significance was set at a threshold of *p*<0.05.

### Exploratory analyses

To determine whether a specific symptom was driving the observed effects, exploratory follow-up analyses were conducted on significant models using scores for the re-experiencing, avoidance, and hyperarousal symptoms. Additional Bonferroni correction was applied to the original corrected threshold for significance to account for the number of symptoms studied for the ROIs (*p*=1.92×10^-3^/3=6.41×10^-4^) and for the VBM maps (*pFWEc*=0.05/3=0.017).

## Results

### Participant characteristics

Demographic details are summarized in Table 1. Participants with chronic pain were not statistically different from the HC group in terms of age, sex and scanning site distributions. However, they reported more severe PTSS, depression, and anxiety than the HC group.

### ROI analyses

Table 2 summarizes the results of all tested statistical models. All models, except those for the left and right pallidum and the left thalamus, were significant (*p*<1.92×10^-3^). Of those, the group-by-trauma interaction was significantly associated with variations in grey matter volume in the left and right putamen, the left MCC, and the right posterior insula (see Figure 1); this was also in the context of significant direct effects of PTSS severity on the left and right putamen. The first set of moderation analyses using PCL-C total score as moderator, indicated that HCs had significantly larger left and right putamen compared to people with chronic pain, only at high levels of PTSS (but not at low or average levels). In addition, people with chronic pain had significantly larger left MCC and right posterior insula compared to the HC group only at low levels of PTSS (not at average or high levels). The second set of moderation analyses using group as the moderator of the relationship between variations in PCL-C scores and ROIs grey matter volumes, indicated that increasing levels of trauma were significantly associated with larger left and right putamen in the HC group only, and with smaller left MCC and right posterior insula in people with chronic pain only.

**Figure 1.**
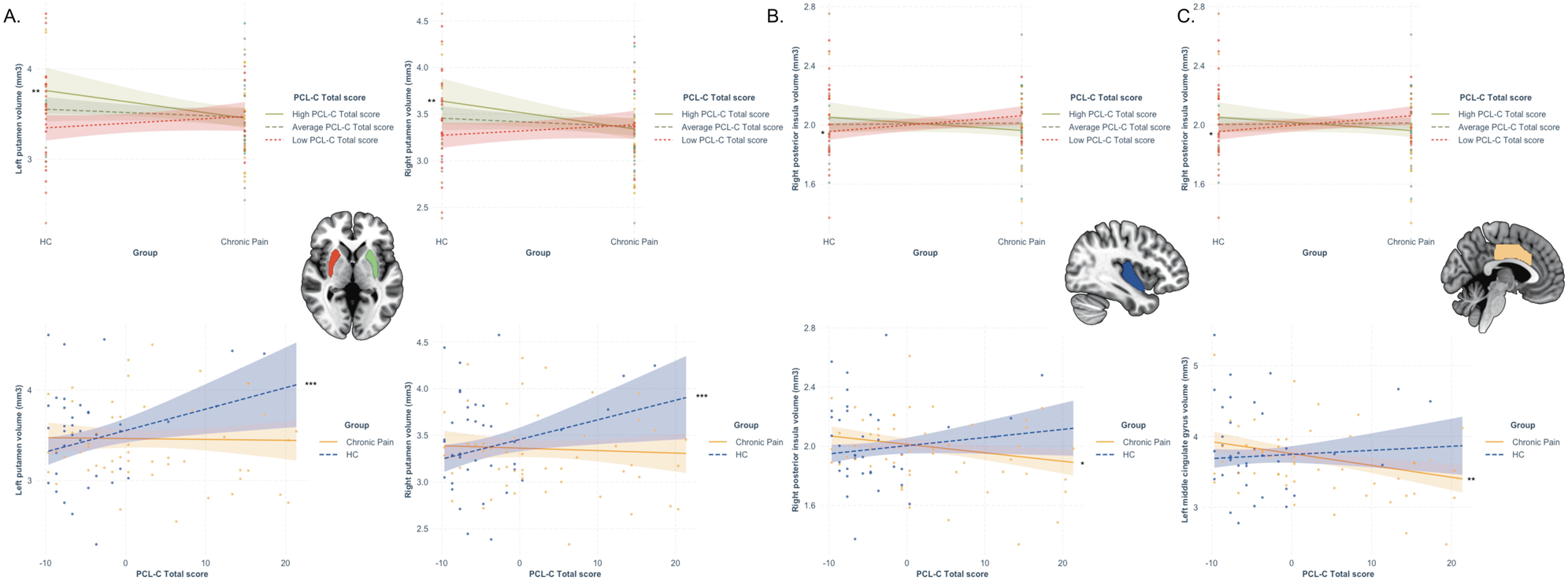
Moderation analyses following significant association between the group-by-PTSS total score interaction term and grey matter volume. The interaction term was significantly associated with variations in grey matter volume of the left and right putamen, the right posterior insula and the left middle cingulum cortex (MCC). (A) When using the PCL-C total score as moderator of the group difference on left and right putamen volumes, analyses indicated that HCs had significantly larger left and right putamen compared to people with chronic pain, only at high levels of PTSS (brown plain lines). When using group as the moderator of the relationship between variations in PCL-C scores and ROIs grey matter volumes, indicated that increasing levels of trauma were significantly associated with larger left and right putamen in the HC group only (blue dashed lines). (B) When using the PCL-C total score as moderator of the group difference on right posterior insula volumes, analyses indicated that people with chronic pain had significantly larger right posterior insula compared to the HC group only at low levels of PTSS (red dashed lines). When using group as the moderator, increasing PCL-C scores were associated with smaller right posterior insula in people with chronic pain only (yellow plain line). (C) When using the PCL-C total score as moderator of the group difference on left MCC volume, analyses indicated that people with chronic pain had significantly larger left MCC than HCs (red dashed line). When using group as the moderator, increasing PCL_C scores were associated with smaller left MCC (yellow plain line). \**p*<0.05; ** *p*<0.01; *** *p*<0.001 Colored band around each line represents 95% confidence intervals.

**Table 2.**
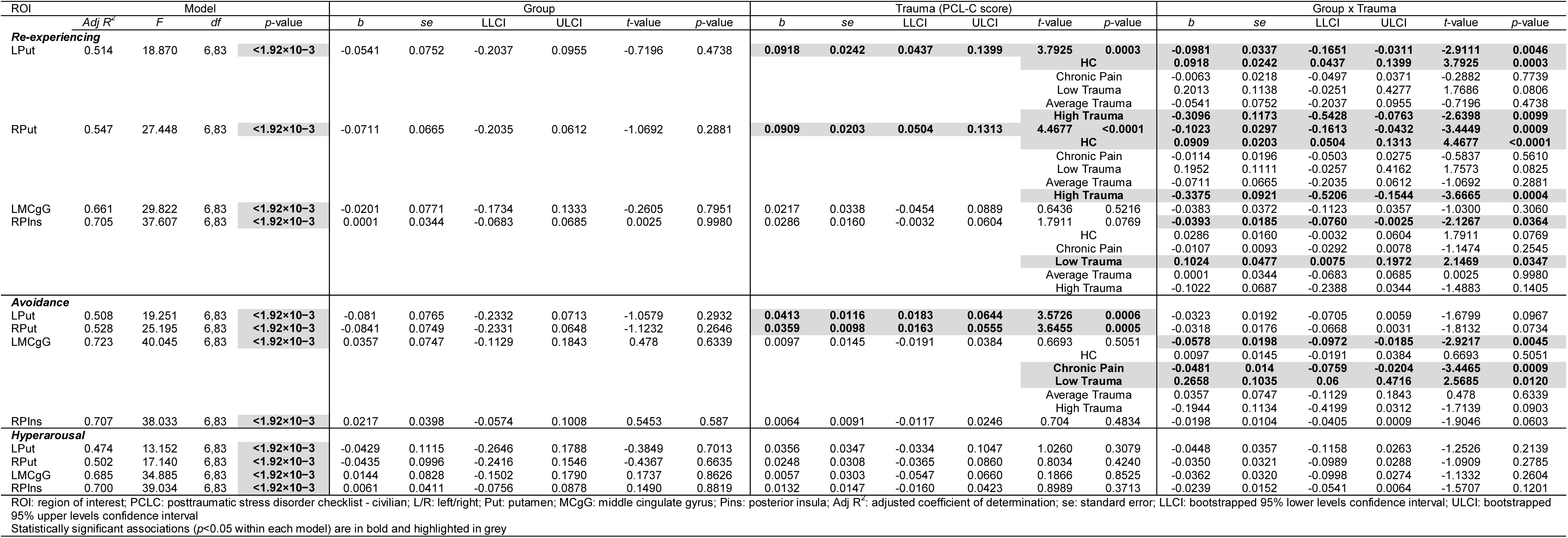
Results of the moderation analyses for all ROIs.

In addition, there was a significant effect of group showing smaller right MFG volume in the chronic pain group relative to the HC group (independent of trauma severity), and a significant increase in volume of the left anterior insula in association with increasing PTSS severity (independent of group).

### Whole-brain analyses

There was no significant association between group, PCL-C total score or their interaction on the whole-brain VBM maps.

### Exploratory analyses

Exploratory analyses were conducted on the ROIs showing a significant association with the group-by-trauma interaction (using the PCL-C total score) above; that is the left and right putamen, the left MCC, and the right posterior insula. Details of these exploratory ROI analyses are presented in Table 3.

**Table 3.**
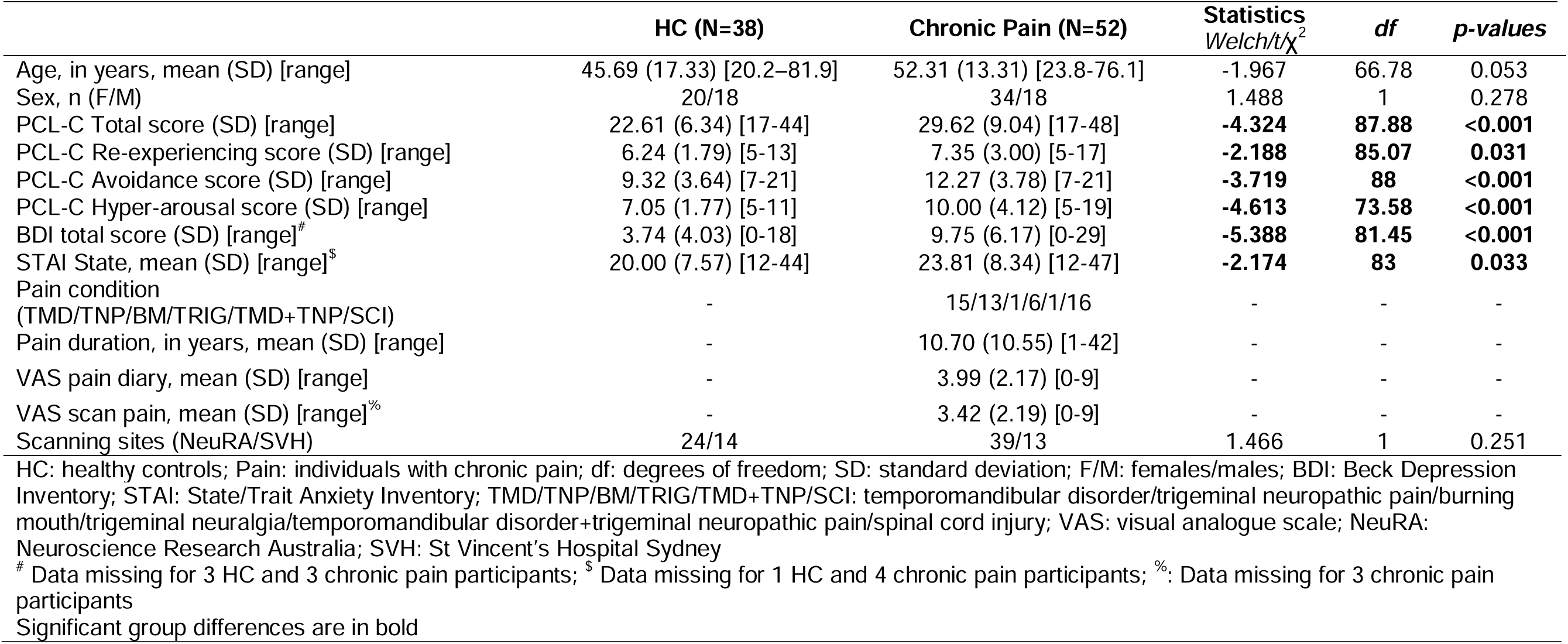
Results of the moderation analyses with separate PTSS symptoms.

#### Re-experiencing

The group-by-trauma interaction was significantly associated with variations in the left and right putamen, and the right posterior insula, but not the left MCC (see Figure 2). The first set of moderation analyses using PCL-C re-experiencing score as a moderator, showed significantly larger left and right putamen at high levels of trauma re-experiencing (but not at low or average levels), and smaller right posterior insula at low levels of trauma re-experiencing (not at average or high levels) in HCs relative to people with chronic pain. The second set of moderation analyses using group as the moderator of the relationship between variations in PCL-C re-experiencing scores and variations in grey matter volume of the ROIs, indicated that larger left and right putamen were significantly associated with increasing levels of re-experiencing in the HC group only.

**Figure 2.**
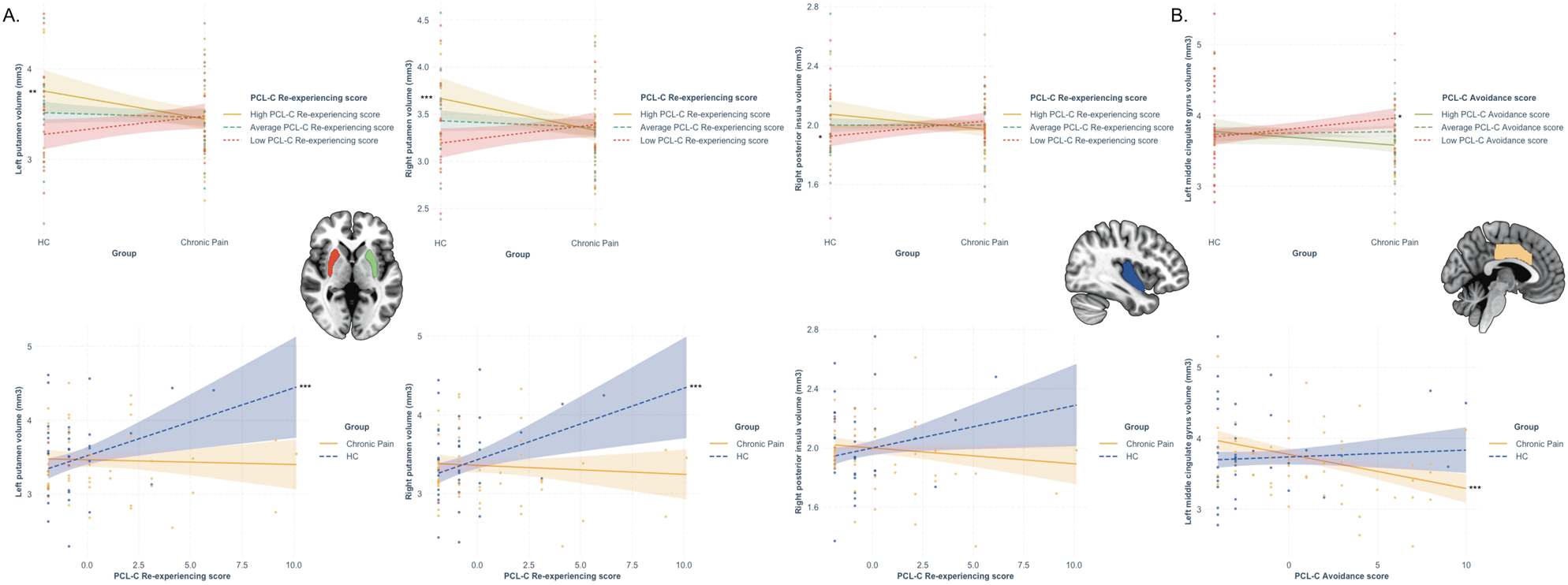
Moderation analyses following significant association between the group-by-PTSS interaction term and grey matter volume. (A) Re-experiencing symptoms significantly moderated the group differences on left and right putamen and right posterior insula. Larger putamen and right posterior insula were evident in HCs compared to people with chronic pain at higher levels of re-experiencing only (brown plain lines). In addition, increasing re-experiencing symptoms were associated with larger left and right putamen only in HCs (blue dashed lines). (B) Avoidance symptoms significantly moderated the group differences on left MCC. Larger left MCC was evident in people with chronic pain compared to HCs at lower levels of avoidance only (red dashed lines). In addition, increasing avoidance symptoms were associated with smaller left MCC only in people with chronic pain (yellow plain lines). * *p*<0.05; ** *p*<0.01; *** *p*<0.001 Colored band around each line represents 95% confidence intervals.

#### Avoidance

Although the associations between left and right putamen and avoidance scores were significant, the group-by-trauma interaction was significantly associated with variations in grey matter volume of the left MCC only (see Figure 2). The first set of moderation analyses using PCL-C avoidance scores as moderator, showed significantly larger left MCC in people with chronic pain relative to the HC group only at low levels of avoidance (but not at average or high levels). The second set of moderation analyses using group as the moderator of the relationship between variations in PCL-C avoidance scores and variations in grey matter volume of the ROIs, indicated that smaller left MCC was significantly associated with increasing levels of avoidance in the chronic pain group only. When exploring associations with whole-brain VBM maps, similar significant association between the group-by-trauma (avoidance scores) and variations in grey matter volume of the bilateral MCC [peak MNI coordinates (-6,-27,42), *k*=1137 voxels, *t(83)*=4.38, *z*=4.14, *pFWEc*=0.017; see Figure 3] was evident. After extraction of the raw signal from the cluster peak, the model was statistically significant (adjusted *R^2^*=0.729, *F*(6,83)=37.027, *p*<0.001) and the interaction was significantly associated with variations in GMV in this cluster (*b*=-0.011, *se*=0.004, *t*=-2.835, *p*=0.006, *95%CI* [-0.018,-0.003]). The first moderation analysis testing PCL-C avoidance as the moderator of associations between groups and GMV (see Figure 3) revealed significant larger GMV in this cluster in the chronic pain group relative to the HC group at low (*b*=0.044, *se*=0.014, *t*=3.186, *p*=0.002, *95%CI* [0.016,0.071]), but not average (*b*<0.001, *se*=0.013, *t*=0.942, *p*=0.942, *95%CI* [-0.024,0.026]) or high levels of PCL-C avoidance (*b*=-0.042, *se*=0.024, *t*=-1.730, *p*=0.087, *95%CI* [-0.090,0.006]). The second moderation analysis testing group as the moderator of associations between PCL-C avoidance and GMV revealed that increasing PCL-C avoidance scores were significantly associated with decreasing GMV in this cluster in the chronic pain (*b*=-0.007, *se*=0.002, *t scores*=-3.760, *p*<0.001, *95%CI* [-0.011,-0.003]) but not the HC group (*b*=0.004, *se*=0.003, *t*=1.080, *p*=0.283, *95%CI* [-0.003,0.010]).

**Figure 3.**
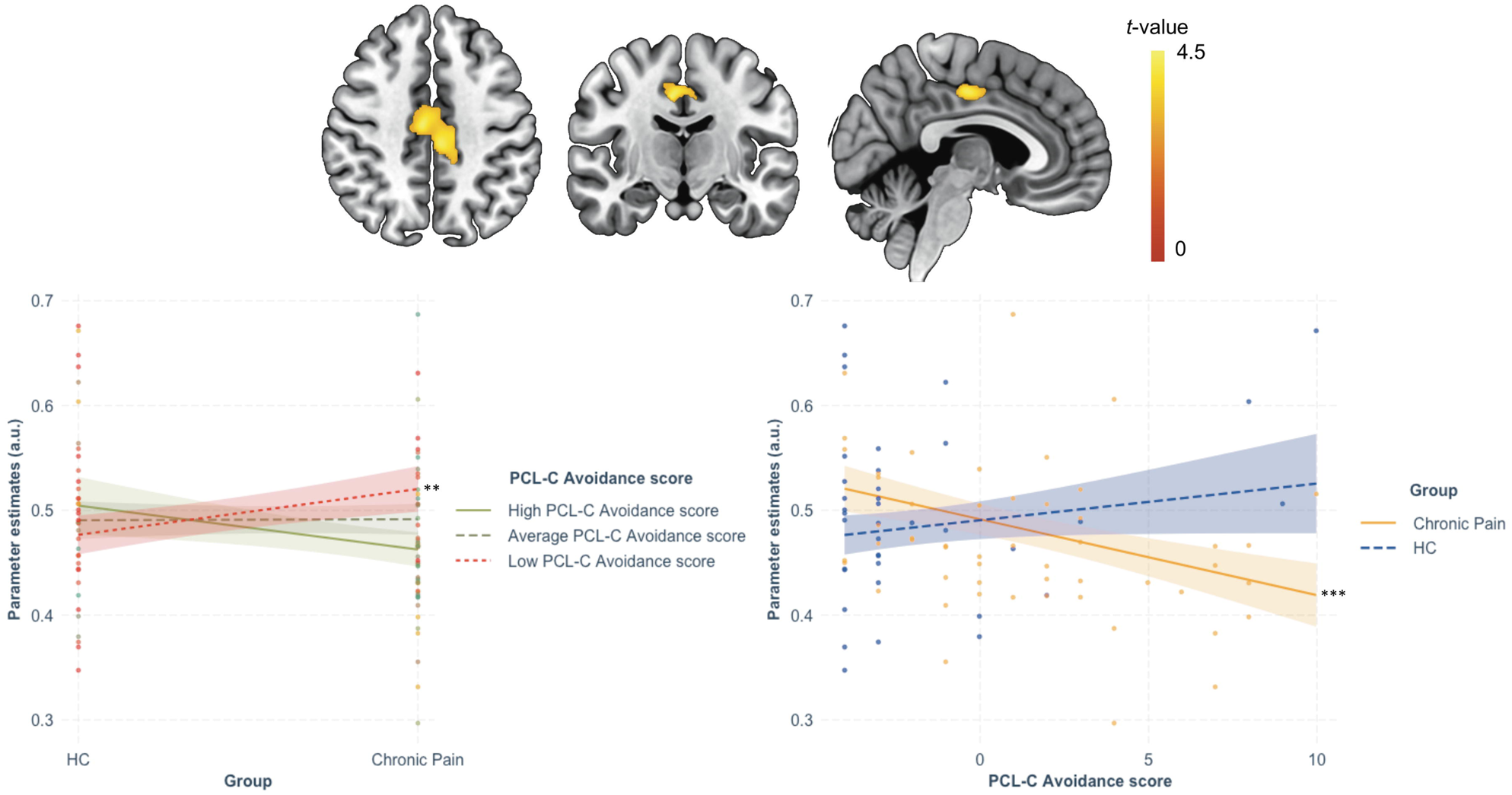
Whole-brain association with the group-by-avoidance interaction. Severity of avoidance moderated the group difference on MCC volume, with the chronic pain group showing larger GMV in this cluster relative to the HC group at low levels of avoidance (left panel; red dashed line). In addition, increasing avoidance severity was associated with smaller GMV in this cluster in people with chronic pain only (right panel; yellow plain line). ** *p*<0.01; *** *p*<0.001 Color-bar represents *t*-statistics. Colored band around each line represents 95% confidence intervals.

#### Hyperarousal

There was no significant effect of group, hyperarousal scores, or group-by-trauma interaction for any of the ROIs or with the whole-brain VBM maps.

## Discussion

This study indicates that PTSS severity impacts brain morphology differently in people with chronic pain, compared to pain-free healthy controls. At higher levels of PTSS, larger putamen and right posterior insula volume were evident in HCs compared to chronic pain, while people with chronic pain had smaller left MCC than HCs at lower levels of PTSS. In addition, increasing PTSS severity was associated with larger putamen volumes in HCs, and with smaller left MCC and right posterior insula in people with chronic pain. Additionally, people with chronic pain exhibited smaller right MFG than HCs, independent of PTSS severity, and PTSS severity was associated with a larger left anterior insula, independent of the group. Exploratory symptom-specific effects were evident in the putamen and right posterior insula for re-experiencing symptoms, and in the left MCC for avoidance symptoms.

Consistent with previous studies of various chronic pain conditions (48), people with chronic pain had smaller right MFG than pain-free HCs. This key part of the dorsolateral prefrontal cortex is critical for executive functions, and for the control and regulation of emotional expression (49). Decreased volume of the dorsolateral prefrontal cortex may reflect a loss of dendrites (50), neurons or lack of neurogenesis in this region that may impact these functions (51). Our findings could also reflect atypical brain aging in people with chronic pain (52), which could be a consequence associated with living with chronic pain. However, this interpretation is speculative given this study did not record indices of cognition, brain function or aging. Other cellular and/or molecular mechanisms not recorded in this study, such as elevated low-grade inflammation, may also have contributed to grey matter alterations in this region, as seen in major depressive disorder (53). Therefore, future studies are warranted, particularly to investigate the relationship between chronic pain and PTSS on cognitive and emotional function in relation to these morphological brain changes.

The severity of PTSS was associated with larger left, but not right, anterior insula and was not associated with grey matter volume alterations in other stress-sensitive regions including the hippocampus, amygdala, mPFC, ACC, and posterior insula. Larger insular volume has been described as a marker of resilience following trauma exposure and associated with trauma severity (54). The present study extends this observation to people with chronic pain. However, it is important to note that most previous studies have not investigated the anterior and posterior insula separately, potentially hindering the identification of more subtle, and specific effects of trauma or chronic pain on more spatially refined subregions. Along with the dorsal ACC, the anterior insula is a core region of the salience network (55) that integrates multi-sensory, exteroceptive and interoceptive stimuli received by the posterior insula (34, 35). Larger anterior insula may thus reflect increased neurogenesis following trauma exposure, potentially helping to better cope with the effects of trauma for those who do not develop fully manifested PTSD. This interpretation will need to be confirmed in future studies.

The severity of PTSS moderated the differences in bilateral putamen, right posterior insula and left MCC volumes between people with chronic pain and pain-free HCs. Larger putamen volume was associated with more severe PTSS, especially re-experiencing symptoms, in pain-free HCs but not for people with chronic pain. There was no overall group difference on these regions, independent of PTSS severity, and no overall association with PTSS severity, independent of group. The putamen is a core dopaminergic hub of the dorsal striatum involved in locomotion and supports learning processes (56). Consistent with a previous study across groups of people exposed (with and without PTSD) and not exposed to trauma, a larger putamen may be related to mechanisms of resilience to trauma in pain-free HCs (57). Larger posterior insula and MCC volumes were evident in people with chronic pain relative to pain-free HCs at lower levels of PTSS, potentially reflecting maladaptive compensatory or recovery effects as a result of developing chronic pain. Moreover, PTSS severity was associated with smaller volumes of these regions in the chronic pain group. In addition to its role in sensorimotor processing (34, 35), the posterior insula is critical for pain perception (32), indicating the detrimental effect of PTSS on morphological integrity of this region in people with chronic pain, potentially contributing to aberrant sensorimotor processing (58). The MCC is critically involved in higher cognitive processes (e.g., cognitive control, conflict-monitoring), body-orientation and movement execution (59), and is key for pain empathy (60). Consistent with similar findings observed in other mental health conditions, including PTSD and psychotic disorders (20, 61), smaller MCC may reflect trauma-related effects on social cognitive networks in people with chronic pain. In the present study, both the ROI and whole-brain analyses reported specific associations between grey matter volume in the MCC and avoidance symptoms, consistent with other PTSD studies (62) and the role of the MCC in pain (63).

This study presents several limitations. First, the sample size was relatively small, which may partly have hindered the discovery of subtle effects. Second, the chronic pain group included a range of different pain conditions and pain was experienced in different body locations (face, lower body), potentially mitigating condition/location-specific effects. However, this approach was the most appropriate to identify large and common brain morphology alterations associated with a shared environmental risk factor (trauma exposure and PTSS) and their interactions. Replication with larger samples is needed to confirm these findings and determine any potential pain condition/location specific effects. Third, it is important to note that pharmacological treatments were not considered in these analyses, pertaining to the presence of pain-free, drug-naïve healthy controls. Additionally, most participants with chronic pain were using a variety of pharmacological treatments. Considering that pharmacological treatments are known to impact brain neurochemistry, morphology, and function (e.g., (64–66)), potential confounds due to pharmacological treatment types and dosages in our results cannot be ruled out.

In conclusion, the present study provides new evidence for the role of PTSS severity in alterations of brain morphology in people with chronic pain. Higher levels of PTSS were associated with larger putamen and right posterior insula in pain-free participants, potentially reflecting mechanisms of resilience to trauma in this group. Higher levels of PTSS, especially avoidance symptoms, were associated with smaller left MCC, a core region of the “pain matrix” with strong links to the somatosensory network and critical for empathy, especially toward pain-related stimuli. This finding is consistent studies investigating similar trauma-related brain regions across other psychiatric conditions. Introducing trauma-related interventions that can influence these brain regions and networks (67), either before or in conjunction with pain-related treatments, may be beneficial to people with chronic pain who report elevated PTSS (68). Replication in larger samples is necessary, especially to confirm the specificity of avoidance symptoms on the MCC. Additionally, further investigation is needed to determine if the observed effects are common to different chronic pain conditions.

## Data Availability

All data produced in the present study are available upon reasonable request to the authors

## Acknowledgments

The authors acknowledge the volunteers who participated in this study, and the assistance of previous students with data collection and entry, and of medical personnel with participant recruitment.

## Data availability

The data that support the findings of this study are available from the corresponding author upon reasonable request.

## Authors contributions

Y.Q. contributed conceptualization, data curation, formal analysis, investigation, methodology, validation, visualization, writing of the original draft, and review and editing. N.H.-S. contributed conceptualization, methodology, and review and editing. N.N.-N. contributed conceptualization, methodology, and review and editing. J.H.M. contributed conceptualization, methodology, supervision, and review and editing. S.M.G. contributed conceptualization, data curation, formal analysis, funding acquisition, investigation, methodology, project administration, resources, supervision, and review and editing.

